# Cross-sectional, interventional, and causal investigation of insulin sensitivity using plasma proteomics in diverse populations

**DOI:** 10.1101/2024.11.09.24317011

**Authors:** Pik Fang Kho, Neil Wary, Daniela Zanetti, Fahim Abbasi, Joshua W. Knowles, Daniel J. Panyard, Katie T. Watson, Laurel Stell, Laura C. Lazzeroni, Stefan Gustafsson, Lars Lind, John R Petrie, Themistocles L. Assimes

**Author notes:** **Corresponding Author:** Themistocles L. Assimes, MD, PhD 3801 Miranda Ave, Palo Alto, CA 94304.

## Abstract

**Background:** We previously reported significant correlations between a direct measure of insulin sensitivity (IS) and blood levels of proteins measured using the Proximity Extension Assay (PEA) in two European cohorts. However, protein correlations with IS within non-European populations, in response to short-term interventions that improve IS, and any causal associations with IS have not yet been established.

**Methods:** We measured 1,470 proteins using the PEA in the plasma of 1,015 research participants at Stanford University who underwent one or more direct measures of IS. Association analyses were carried out with multivariable linear regression within and across Stanford subgroups and within each of the two European cohorts. Association statistics were also meta-analyzed after transformation and harmonization of the two direct measures of IS. Lastly, we performed genome-wide association studies of IS and used genetic instruments of plasma proteins from the UK Biobank to identify candidate causal proteins for IS through Mendelian Randomization (MR) analysis.

**Results:** In age and sex adjusted model, 810 proteins were associated with baseline IS among 652 self-reported European participants in the Stanford cohort at a false discovery rate (FDR) < 0.05. Effect sizes for these proteins were highly correlated with those observed in 122 South Asian, 92 East Asian, 85 Hispanic, and 52 Black/African American persons (r= 0.68 to 0.83, all P≤4.3×10^-113^). Meta-analysis of the full Stanford cohort with the two European cohorts (N=2,945) yielded 247 significant protein associations (FDR < 0.05), with 75 remaining significant after further adjustment for body mass index. In a subset of Stanford participants undergoing insulin sensitizing interventions (N=53 taking thiazolidinediones, N=66 with weight loss), 79.6% of protein level changes were directionally consistent with the respective baseline association (observed/expected p=6.7x10^-16^). MR analyses identified eight candidate causal proteins for IS, among which were SELE and ASGR1, proteins with established drug targets currently under investigation.

**Conclusion:** Plasma proteins measured using the PEA provide a robust signature for IS across diverse populations and after short-term insulin sensitizing interventions highlighting their potential value as universal biomarkers of insulin resistance. A small subset of markers provided insights into potential causal molecular mechanisms and therapeutic targets.

**Highlights:**

- Insulin sensitivity-related plasma proteins are consistent across diverse populations.
- Protein changes from interventions align with baseline, aiding insulin sensitivity tracking.
- SELE and ASGR1 are potential targets for insulin sensitivity.

## Introduction

Insulin resistance (IR) is a physiological state characterized by impaired insulin-stimulated glucose uptake, leading to hyperinsulinemia, aberrant glucose homeostasis, elevated blood pressure, atherogenic dyslipidemia, and disorders of coagulation, inflammation and endothelial function [1, 2]. Collectively, these co-occurring abnormalities are often referred to as the insulin resistance syndrome, and they substantially increase the risk of type 2 diabetes, atherosclerotic cardiovascular disease (CVD), metabolic associated fatty liver disease, obstructive sleep apnea, polycystic ovary syndrome and certain cancers [3].

The molecular mechanisms underlying the development and progression of IR remain incompletely characterized yet, understanding these mechanisms is crucial for developing new therapies. Accurate quantification of insulin sensitivity (IS) is essential, but current gold-standard methods, including the hyperinsulinemic-euglycemic clamp (HEC) and the insulin suppression test (IST) [4], are labor-intensive and invasive [4, 5]. Proxy measures of IS exist and, while these measures are much easier to obtain, they correlate only moderately with the two gold standard measures [6–11]. This situation underscores the need for alternative approaches considering the high cost and complex nature of acquiring a gold standard measure.

Using the proximity extension assay (PEA)[12], we previously identified a protein signature that correlates highly with the M-value derived from HEC test [13]. We found that LASSO derived models that included 34 to 67 of the measured proteins increased the absolute variance explained of the HEC-derived M-value by as much as 24% over clinical variables alone. While these results are promising, further improvements of this signature are needed before it can serve as an effective surrogate for a direct test. Here, we build on our prior work by successfully measuring close to 1500 proteins using an expanded PEA panel in a more diverse cohort of research participants who underwent one or more ISTs at Stanford University. The new data allowed us to expand discovery of proteomic associations, assess consistency of these associations across major self-reported racial/ethnic groups and after insulin sensitizing interventions, and enabled the first extensive MR study of plasma proteomic correlates of direct measures of IS.

## Methods

### Study Cohorts

We utilized data from three cohorts: the Relationship between Insulin Sensitivity and Cardiovascular Disease (RISC), Uppsala Longitudinal Study of Adult Men (ULSAM), and Stanford studies of insulin resistance (IR). The details and methodology of RISC and ULSAM have been previously described [14, 15]. Briefly, the RISC study was a multicenter, prospective observational cohort that spanned 19 centers in 14 European countries [15]. It involved up to 1,030 participants with protein data and 786 participants with genotype data, all healthy individuals aged between 30 and 60, aiming at predicting cardiovascular disease risk based on insulin sensitivity during a 3 to 10-year follow-up [15]. The ULSAM study is an ongoing longitudinal epidemiological study of up to 900 participants with protein data and 1,030 with genotype data, consisting of men born between 1920 and 1924 in Uppsala County, Sweden [14], with HEC performed at age 70 in the early 1990s. These participants underwent interviews and examinations at the ages of 50, 60, 70, 77, 82, 88 and 93 [16, 17].

The Stanford studies of IR included 1,015 volunteers who participated in 14 research protocols approved by the Institutional Review Board evaluating the role of insulin resistance in human disease between April 2001 and January 2020 [18–21]. Participants were recruited through print and social medial advertisements and provided written informed consent. The participants were generally in good health and ranged in age from 18 to 80 years. This cohort was racially/ethnically diverse by self-report, and included 652 White/European, 122 South Asian, 92 East Asian, 85 Hispanic, and 52 Black/African American. The participants with diabetes were either receiving treatment with one or more glucose lowering medications for management of hyperglycemia or had fasting glucose level ≥126 mg/dL on more than one occasion.

A subset of participants in the Stanford studies of IR underwent insulin sensitizing interventions including taking thiazolidinediones (TZDs) or adopting a diet protocol leading to significant weight loss [22–24]. In these protocols, participants underwent a second IST to assess improvements in IS. From these studies, we included the subset of subjects who had ≥ 25% decrease in steady-state plasma glucose (SSPG) concentration (improvement in IS) following TZD treatment (N=53) or attained ≥ 5% weight loss and ≥ 20% decrease in SSPG concentration with diet (N=66). These thresholds were chosen to ensure the detection of meaningful and marked physiological changes.

Among TZD subgroup, 45 were treated with pioglitazone, and 8 were treated with rosiglitazone [22–24]. Participants underwent baseline measurements prior to initiating treatment with pioglitazone or rosiglitazone. Pioglitazone treatment was started at a dose of 15 mg daily for the first 2 weeks. At the end of 2 weeks, pioglitazone dose was increased to 30 mg daily for the next 2 weeks, followed by 45 mg daily for 8 weeks. Rosiglitazone treatment was started at a dose of 4 mg daily for weeks, followed by 4 mg twice daily for 8 weeks. All participants were instructed to maintain their usual physical activity level for the duration of the study. Following treatment with pioglitazone or rosiglitazone, all baseline measurements were repeated.

Among the 66 participants in the weight loss subgroup [22, 25, 26], 45 achieved weight loss through a reduced-calorie diet (RCD), 12 through a combination of RCD and liraglutide, and 9 underwent bariatric surgery. Participants undergoing weight loss through a RCD were provided dietary instructions by a registered dietitian, aiming to achieve a weekly weight loss of 0.5 kg. Participants were seen every 1 to 2 weeks for weight measurements and dietary advice. In the liraglutide group (N = 12), in addition to RCD, participants began treatment with liraglutide at a daily dose of 0.6 mg. The liraglutide dose was increased by 0.6 mg each week, reaching a maximum dose of 1.8 mg. In the bariatric surgery arm, participants underwent weight loss bariatric surgery. The period of weight loss was 3 months. Upon completing the weight loss phase, participants were instructed on a weight maintenance diet. After maintaining a stable weight for 2 weeks, all baseline measurements were repeated.

### Measurement of Insulin Sensitivity (IS)

IS was assessed directly across the cohorts using two different established but highly correlated methods. In RISC and ULSAM cohorts [14, 15], IS was obtained through the HEC [27]. This method involves administration of a constant infusion of insulin to achieve hyperinsulinemia and a variable infusion of glucose to maintain euglycemia. During the test, the rate of glucose infusion (M value) needed to maintain euglycemia serves as a measure of insulin sensitivity, where a higher M value indicates higher IS. In the Stanford cohort, IS was measured as the SSPG, obtained via the IST [28]. This method involves administering a constant infusion of insulin, octreotide, and glucose. During the test, the magnitude of SSPG concentration achieved under a similar level of hyperinsulinemia approximately 3 hours after the start of the infusions serves as the measure of IS, where a lower SSPG concentration indicates higher IS [29, 30]. These measures are considered “gold standard” measures of IS [31, 32], and are highly correlated with each other [5, 33]. To harmonize these measures where needed, SSPG values were converted to M values using established formulae [5].

### Proteomic profiling

Blood samples were collected after an overnight fast on the day of the IST from all participants in the Stanford studies of IR. Following collection, plasma was immediately separated, aliquoted, and stored at -80 Celsius. All samples were not thawed before the measurements. Proteins were quantified by Psomagen, a certified provider of Olink Proteomics services, using the initial Olink Explore release that included four disease and biological process focused 384-plex panels of proteins: cardiometabolic, inflammation, oncology, and neurology [12]. All protein values, quantified as Normalized Protein eXpression (NPX), were transformed within Olink’s MyData Cloud Software using a log2 scale across studies. This transformation ensures consistency in the comparison of effect estimates and the combination of these estimates in meta-analysis.

A total of 1,144 plasma samples were sent for proteomic profiling including the subset of persons with one sample before and one sample after intervention. To minimize batch effects, participants with two samples before and after interventions were plated on the same plate.

After quality control (QC) procedures, 1,134 participants and 1,470 proteins were retained for analysis, with 1,015 participants having baseline phenotypic data and 119 having post-intervention phenotypic data (**Supplementary Table 1**). QC procedures are described in detail in the supplementary file.

Proteomic measurements and QC for the RISC and ULSAM is extensively described elsewhere [13]. In brief, 787 proteins were measured and passed QC in these two cohorts using nine Olink Target panels. All proteins were also available and passed QC in our new data generated in the Stanford studies of IR using the first commercially available Explore panel.

### Genotyping

We genotyped 937 samples from the Stanford cohort using the Axiom^TM^ Precision Medicine Diversity Array by Applied Biosystem and imputed genotypes to the TOPMed reference panel [34–36]. Genotyping and imputation for the RISC and ULSAM cohorts are described in detail elsewhere [37]. Our genome-wide association studies (GWAS) and Mendelian Randomization (MR) analyses were restricted to European participants and included 571 out of 596 samples that passed quality control (QC), of which 512 also had IS data. QC procedures are described in detail in the supplementary file. We applied the same QC as the Stanford cohort, with 767 and 1,030 European samples passing QC in RISC and ULSAM, respectively.

### Statistical Analysis

#### Associations of plasma proteins and insulin sensitivity in cross-sectional studies

We ran two regression models to identify plasma proteins linked to SSPG in the Stanford studies of IR while accounting for potential confounders. The first model controlled for age, sex, and study protocol, while the second additionally controlled for BMI, a primary determinant of IS. We applied both false discovery rate (FDR) and Bonferroni corrections to p-values to determine significance in the presence of multiple testing. The Bonferroni correction, while being more stringent, controlled for the number of proteins tested, whereas the FDR correction, while being more lenient, controlled for false positives while allowing for the correlation between proteins.

Proteins that showed significant associations in White/European participants were subsequently tested for their associations with SSPG in each of the other self-identified race/ethnicity (SIRE) groups. To assess the generalizability of these associations across these groups, we determined the Pearson correlation between the beta effect estimates in white/European persons with that observed in each of the four non-European subgroups. Lastly, we carried out a random-effect meta-analysis across the three cohorts to extend discovery of associations with IS for the 802 protein measures available in all three cohorts. For this analysis, we used the Restricted Maximum Likelihood (REML) method in the “metafor” package to account for between-study heterogeneity [38]. To accomplish this task, we first conducted linear regressions within RISC and ULSAM. Then, we repeated regression analyses in all participants of the Stanford studies of IR after transforming the SSPG to an M-value and additionally controlling for self-reported race/ethnicity. Once again, we applied both FDR and Bonferroni corrections to p-values to address multiple testing.

#### Evaluation of changes in protein levels after insulin sensitizing interventions

Proteins with significant associations at FDR < 0.05 from the BMI-unadjusted meta-analysis were selected for further evaluation within the insulin sensitizing intervention subgroup who underwent a second proteomic profiling after the intervention. In this subset, we used linear regression to assess the relationship between changes in protein levels and corresponding changes in SSPG concentration between the first and the second IST. We compared the direction of the effect sizes observed in the meta-analysis of cross-sectional studies (using M values) with those from the Stanford Intervention Studies (using SSPG concentration as the measure of IS). Given the strong inverse correlation between M values and SSPG concentration, this comparison allowed us to assess the consistency of the protein associations across different studies. All analyses were repeated after further adjusting for BMI.

#### Causal relationships between candidate proteins and insulin sensitivity

To investigate the causal relationships between 247 candidate proteins and IS, we employed two-sample MR analysis [39]. Prior to the MR analysis, we conducted a GWAS of insulin sensitivity (measured as M value) within European participants of each cohort using REGENIE [40]. M values were inverse rank transformed prior to GWAS. REGENIE’s two-step regression approach allowed us to account for relatedness and population structure, with age, sex, protocol number and top 10 principal components included as covariates [40]. We subsequently integrated summary statistics from the RISC, ULSAM and Stanford cohorts (N = 2,309) using a fixed-effect model in METAL software [41].

We used the summary-level data from the UK Biobank to identify instrumental variables for 247 candidate proteins [42]. These instrumental variables were primarily cis-protein quantitative trait loci (cis-pQTLs), selected based on their strong associations with protein levels (P-value < 5x10^-8^) and their proximity within +/-500kb of the corresponding protein-coding genes. Where cis-pQTLs were not available, trans-pQTLs were used. To ensure independence of the instrumental variables, we clumped SNPs using a 10Mb linkage disequilibrium (LD) window with an r^2^ threshold of < 0.001, using the 1000 Genomes European Project [43] as the reference panel. Eight proteins located on chrX (ACE2, ADGRG2, F9, IDS, IL13RA1, ITM2A, LAMP2, and VEGFD) were excluded from MR analysis, where we focused solely on autosomal SNPs. We performed Wald-type ratio and inverse variance weighted (IVW) analyses as MR primary analyses, depending on whether one or multiple SNPs were available as instrumental variables. We also performed sensitivity analyses to evaluate the robustness and validity of the results while accommodating the presence of different levels of pleiotropy. These sensitivity analyses included weighted median regression, weighted mode regression, and MR-Egger regression. All MR primary and sensitivity analyses were performed using the “TwoSampleMR” package in R [39]. For each protein, we calculated the proportion of variance (R^2^) explained by instrumental variables using the following formula [44]:

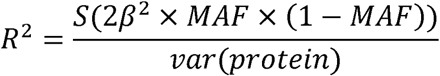

where *β* is the effect size of a given SNP on the plasma protein level, and MAF is the minor allele frequency of the SNP, is the standard error of the effect size, and N is the total sample size.

## Results

Key characteristics of the 1,015 Stanford participants included in our association analyses are summarized in **Table 1**. Approximately 35% of the sample was of non-White/European SIRE group. There were no significant differences in sex and fasting plasma glucose distributions across race/ethnic groups (**Table 1**). However, significant variations were observed in age with South Asians being the youngest and White/Europeans the oldest (**Table 1**). BMI also showed significant differences across races with South Asians had the lowest mean BMI whereas Black/African Americans had the highest (**Table 1**). SSPG levels also differed significantly with Hispanics having the highest and White/Europeans having the lowest (**Table 1**). Accordingly, converted M values varied equivalently but in the expected opposite direction to the SSPG levels across groups.

**Table 1.** Baseline characteristics of participants profiled from the Stanford Studies of Insulin Resistance stratified by self-reported race ethnicity.

| SIRE | Black/African American |  | East Asian |  | Hispanic |  | South Asian |  | Two or more races |  | White/European |  | P value |
| --- | --- | --- | --- | --- | --- | --- | --- | --- | --- | --- | --- | --- | --- |
| n | 52 |  | 92 |  | 85 |  | 122 |  | 12 |  | 652 |  |  |
| sex (%) |  |  |  |  |  |  |  |  |  |  |  |  |  |
| Female | 29 | 55.8 | 43 | 46.7 | 43 | 50.6 | 71 | 58.2 | 6 | 50 | 356 | 54.6 | 0.63 |
| Male | 23 | 44.2 | 49 | 53.3 | 42 | 49.4 | 51 | 41.8 | 6 | 50 | 296 | 45.4 |  |
| age (mean (SD)) | 46.52 | 10.11 | 49.26 | 10.3 | 49.02 | 10.38 | 42.8 | 9.7 | 48.17 | 9.67 | 52.42 | 9.75 | <0.001 |
| BMI (mean (SD)) | 32.63 | 7 | 28.73 | 3.73 | 31.75 | 5.71 | 28.24 | 3.84 | 31.78 | 3.58 | 29.61 | 4.79 | <0.001 |
| Fasting plasma glucose (mean (SD)) | 97.42 | 16.77 | 101.1 | 16.85 | 100.86 | 13.05 | 97.98 | 15.35 | 93.33 | 9.15 | 100.13 | 15.91 | 0.29 |
| SSPG (mean (SD)) | 162.06 | 76.46 | 170.93 | 63.8 | 187.29 | 84.46 | 169.33 | 72.67 | 151.25 | 56.93 | 149.8 | 73.28 | <0.001 |
| Converted M (mean (SD)) | 5.23 | 3.17 | 4.61 | 2.65 | 4.49 | 3.37 | 4.85 | 2.91 | 5.31 | 2.85 | 5.76 | 3.23 | <0.001 |
| SIRE: self-identified race/ethnic groups; n: Sample size; SD: standard deviation; SSPG: steady-state plasma glucose. Data are mean ± standard deviation (SD) (range) unless otherwise noted. Means were compared by paired sample t test and proportions by chi-square test. Converted M = $18.309 \times \exp(-0.009 \times \text{SSPG})$ | | | | | | | | | | | | | |

We identified 810 proteins associated with SSPG concentration in the Stanford White/European participants at FDR < 0.05, with 395 passing Bonferroni correction (**Supplementary Figure 1; Supplementary Table 2**). Adjusting for BMI reduced this number to 570 at FDR < 0.05, with 200 passing Bonferroni correction (**Supplementary Figure 1; Supplementary Table 3**). Pearson correlation analysis documented a moderately high correlation between the effect sizes in White/European participants and each of the four other major SIRE groups (r=0.68 to 0.83, all P<= 4.3×10^-113^) for the 810 associated proteins that were associated with SSPG concentration in the Stanford White/European at an FDR < 0.05. The correlation coefficients increased when considering only the 395 proteins that passed Bonferroni correction (r=0.79 to 0.89 all P<=7.3×10^-85^) (**Figure 1**). These correlations were consistent with analogous analyses using significantly associated proteins in models that further adjusted for BMI.

**Figure 1.**
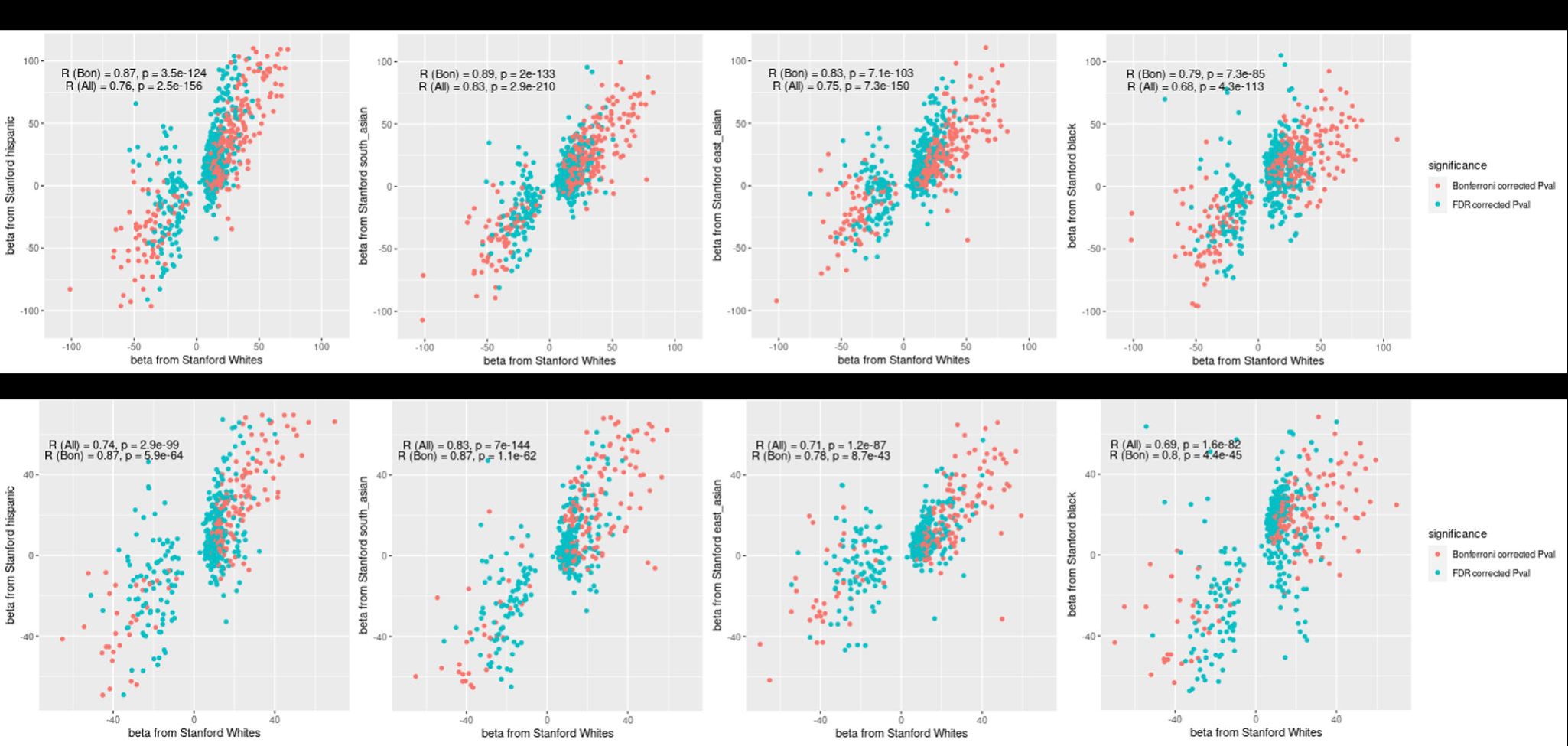
Comparison of regression coefficients for different populations in (a) BMI-unadjusted and (b) BMI-adjusted models, using White/Europeans in the Stanford study as reference. The comparisons are based on the proteins associated with SSPG in Whites. Pearson correlations between each pair of populations are also reported.

A total of 778 unique protein measures passed QC in all three cohorts (RISC, ULSAM, and Stanford). Pearson correlation analysis revealed a strong correlation of effect sizes (r = 0.89) between the 164 proteins that were nominally significant in both the meta-analysis of the RISC and ULSAM cohorts (P<0.05) and the multi-ethnic Stanford studies of IR (P<0.05), which increased to 0.94 restricting to the 102 and 38 proteins passing FDR and Bonferroni corrections respectively(**Supplementary Figure 2a; Supplementary Tables 2 and 4**). When we next integrated all three sets of data (Stanford n=1,015, RISC n=1,030, and ULSAM n=900) through meta-analysis, we identified 247 unique candidate plasma proteins at an FDR < 0.05, with 49 remaining significant after BMI adjustment (**Figure 2**; **Supplementary Table 5**). Of these, 142 of the BMI-unadjusted and 29 of the BMI-adjusted significantly associated proteins were measured in all cohorts while 106 and 21, respectively were only measured in the Stanford cohorts.

**Figure 2.**
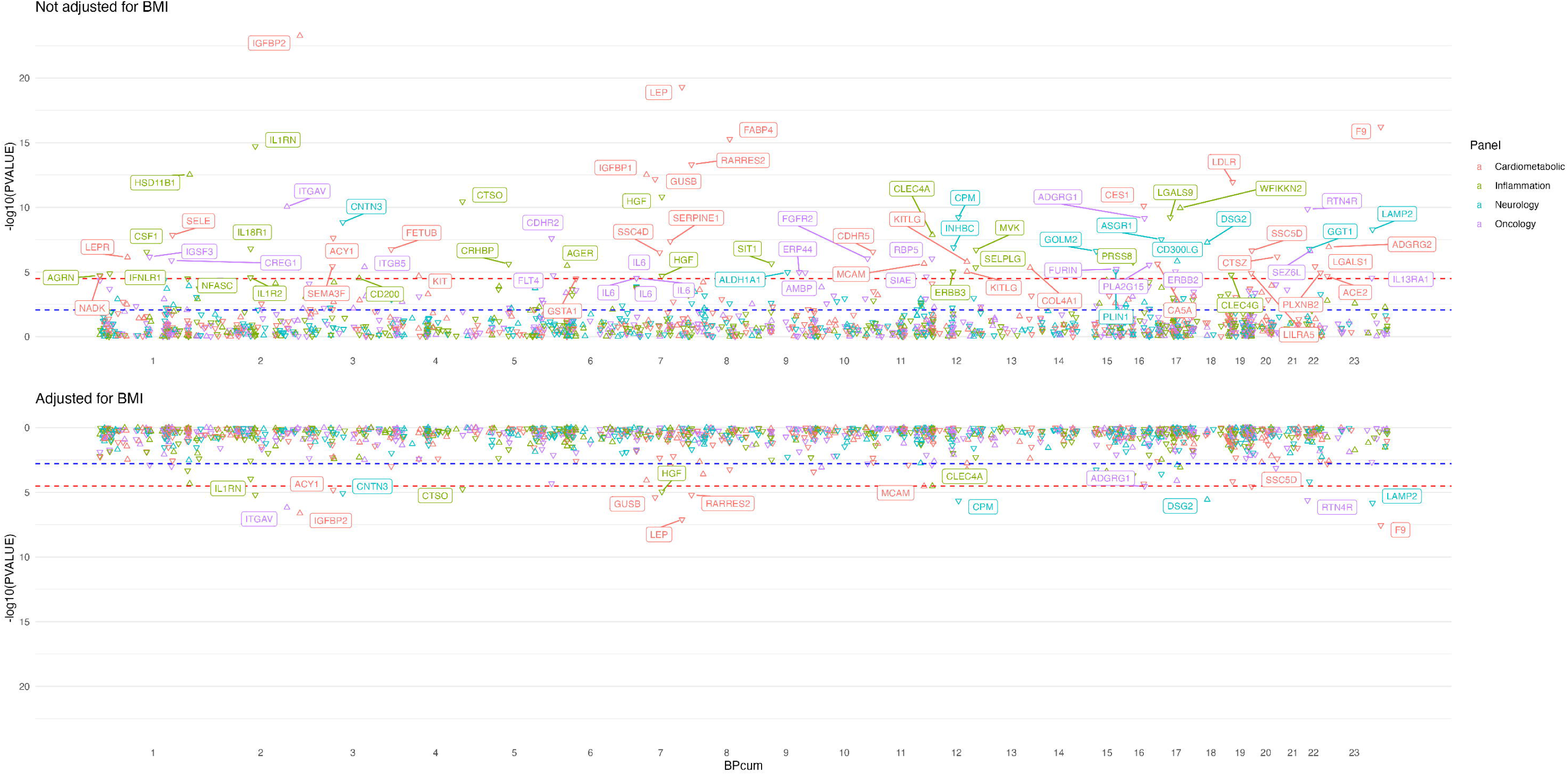
Manhattan plot of meta-analysis combining cross-sectional studies of plasma proteins and M values from RISC, ULSAM and Stanford. Top panel shows the BMI-unadjusted model and the bottom panel shows the BMI-adjusted model. Proteins are color-coded according to their Olink panel. Red dotted lines indicate Bonferroni-corrected P-values (P < 0.05/1,471 proteins tested = 3.4×10^-5^) while blue dotted lines indicate FDR-corrected P-values. Proteins that passed Bonferroni correction are annotated, and the direction of effect of each protein on M values is indicated by the direction of the triangle.

Next, we compared the baseline associations with IS for the 247 significant proteins identified from the meta-analysis levels with the change in SSPG levels (delta SSPG) of these same proteins occurring after TZD or weight lost intervention among the Stanford participants (**Supplementary Tables 6-7**). We compared the effect sizes (betas) of baseline protein levels with M values with those from changes in protein levels with improvements in SSPG after interventions. We observed a 79.6% concordance between the baseline direction of effect and the expected direction of effect for the change (x^2^, p=6.7x10-^16^, **Figure 3**). Concordance proportions were lower within each intervention subgroup, with 64.7% concordance in TZD treatment subgroup (x^2^ p=6.3x10^-3^) and 67.8% in weight loss subgroup (x^2^ p=6.7x10^-6^, **Figure 3**, Supplementary Table 8).

**Figure 3.**
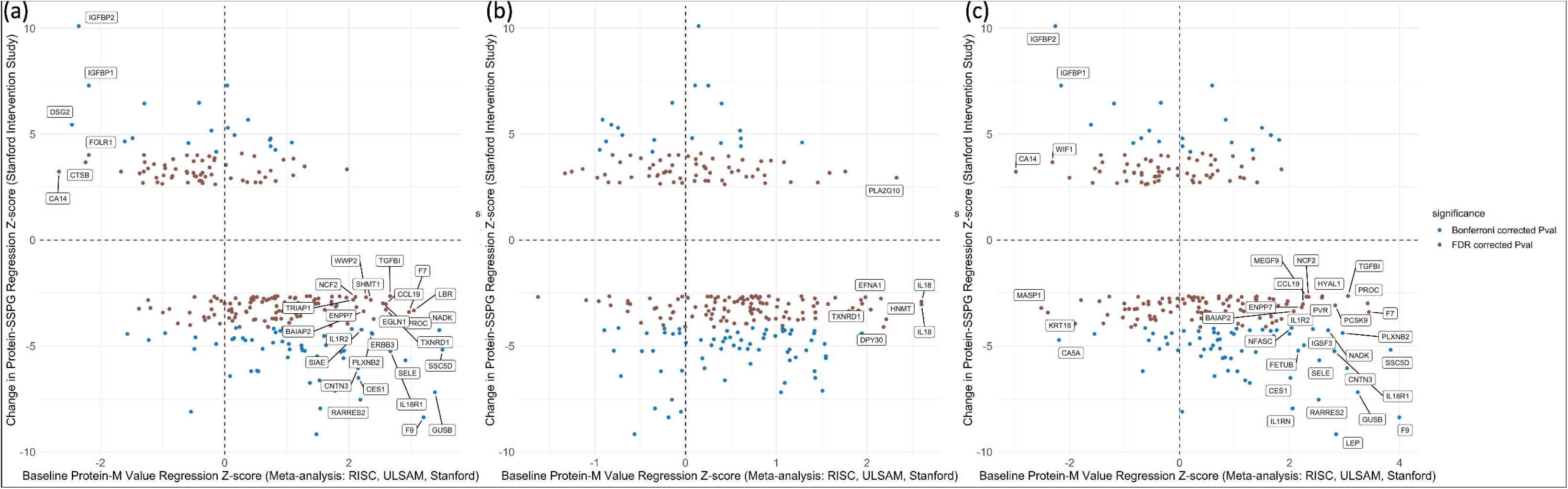
Correlation of regression Z-score between meta-analysis of cross-sectional studies and Stanford intervention study of (a) all participants, (b) those who underwent TZD intervention and (c) those who underwent weight loss intervention, using 255 candidate proteins identified in the meta-analysis of cross-sectional studies at FDR < 0.05. Insulin sensitivity was measured as M values in meta-analysis of cross-sectional studies, and as SSPG in Stanford Intervention study. Proteins with significant associations at P < 0.05 were annotated.

Lastly, we explored the causal relationships of these 247 candidate proteins of IS using two-sample MR analysis. Leveraging cis-pQTLs as instrumental variables, we identified twelve proteins with potential causal associations with M values in MR primary analysis at P < 0.05 (**Figure 4**, **Supplementary Table 9**). Ten of twelve proteins were further corroborated by at least one sensitivity analysis when available (**Figure 4**, **Supplementary Table 9**). CDHR5, CES3, INHBC, PTPRN2 MASP1 and MVK were positively correlated (greater insulin sensitivity) while IL10RB, ASGR1, SELE, and SHMT1 were negatively correlated (lower insulin sensitivity) with M values. Among the proteins analyzed, PLXNB2 had the strongest instrumental variable, with its cis-pQTLs explaining 24.2 % of the variance in plasma PLXNB2 levels (**Supplementary Table 9**). Despite its strong instruments, <u>PLXNB2</u> did not achieve statistical significance in the MR primary analysis. For significantly associated proteins (p<0.05) in the primary MR, the proportion of the variance explained by the cis-pQTLs used as instruments ranged from 0.32% to 13.3% (**Supplementary Table 8**). For the 22 proteins without available cis-pQTLs, we leveraged trans-pQTLs as instrumental variables. However, none of these proteins showed significant associations with M values in the MR analysis at P < 0.05.

**Figure 4.**
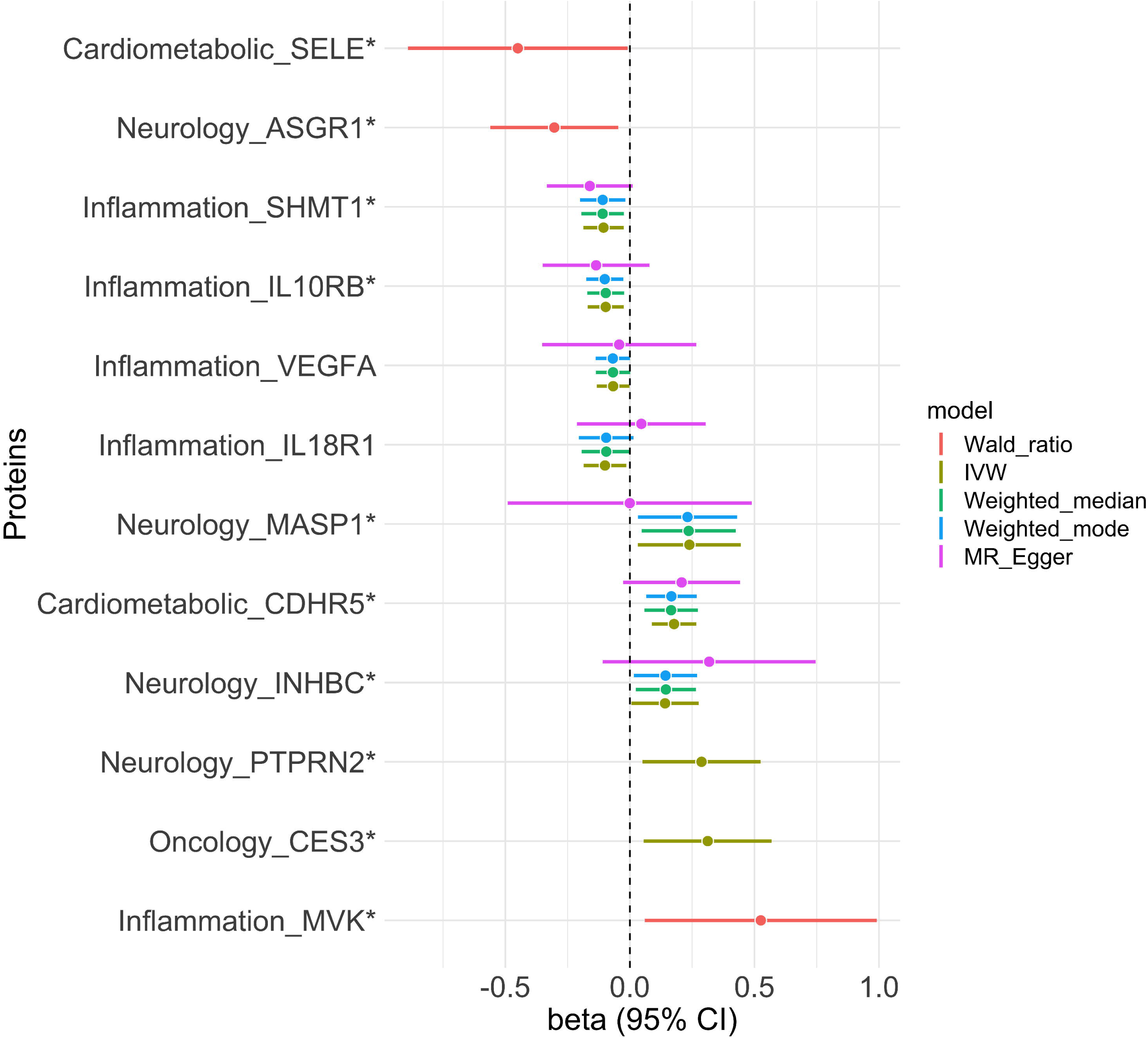
Mendelian randomization analysis of cis-pQTLs only of candidate causal proteins on M values. The proteins shown are those with evidence of causal associations in MR primary analyses at P < 0.05. Candidate causal proteins supported by at least one sensitivity analysis (where available) at P < 0.05 are marked with an asterisk.

## Discussion

We report the first large-scale investigation of the proteomic determinants of direct measures of insulin sensitivity (IS) across diverse populations and among subjects undergoing randomized interventions. We identified novel proteins associated with IS and demonstrate a high consistency of the magnitude and direction of associations across multiple cohorts and multiple ancestral groups, highlighting the reproducibility and the generalizability of the associations. These proteins also track with changes in IS following insulin sensitizing interventions, suggesting their potential utility in monitoring therapeutic effectiveness. Lastly, instrumental variable analyses prioritize eight proteins with potential causal effects on IS including at least two, ASGRI and SELE, that are targets of existing drugs.

Our previous work profiled participants exclusively of European ancestry within the RISC and ULSAM cohorts based in Europe using nine Olink 96-plex Target panels that combine the PEA assay technology with an integrated microfluidic chip and a qPCR based data readout [13]. Here, we leveraged the first four 384-plex high-throughput panels of the Explore platform integrating a next generation sequencing data readout to profile over 1,000 subjects of diverse SIRE living in the San Francisco Bay Area. A large majority (778/829, 94%) of the proteins that were previously successfully profiled in RISC and ULSAM also passed QC in the Stanford cohorts using the Explore platform. This design allowed for a comparison as well as a harmonization and meta-analysis of results across all three cohorts.

We found over half of the proteins (810/1,470, 55%) measured among the SIRE white/Europeans in the Stanford cohorts to be associated with SSPG at a conventional FDR threshold with about one half of these (or 25% overall) meeting the more stringent Bonferroni threshold of significance. This remarkable degree of significance was validated by comparing protein-associations available in all cohorts which demonstrated not only a high rate of replication but also a high correlation of effect sizes. About one in five proteins remained significant among the subset of proteins available in all cohorts after meta-analysis (142/778, or 18%) while about one in six measured only in the Stanford cohorts was significant (106/693, or 15%). The number and the fraction of significantly associated proteins dramatically reduced after adjusting for BMI (29/778, or 3.7% and 21/693, or 3%), reinforcing the well-established importance of adiposity as a primary driver of IS. Overall, these findings likely reflect a combination of factors including the presence of a strong proteomic signature in the plasma for a key physiologic state such as IS, a biased selection of measured proteins matching the traits most relevant to IS including adiposity, dyslipidemia and hypertension (e.g. cardiometabolic and inflammation panels), and a modest but pervasive correlation of protein levels selected for measurement overall.

We demonstrate the potential utility of using a proteomic signature to more optimally quantify and track changes in IS in the setting of clinical care through our analyses of nonwhite/European populations and changes in protein levels after interventions leading to a substantially improved IS. First, we demonstrated moderate-to-high concordance of associations of proteins with IS across race/ethnic groups in our multi-ancestry Stanford cross-sectional study. However, some differences were noted between White/European and Black/African American participants. The differences could be due to statistical power limitations, as we have less than one tenth Black/African Americans in this study compared to white/Europeans. Nonetheless, the overall consistency suggests these associations can potentially be extrapolated across populations. Additionally, we must not discount the possibility that true differences in protein-IS associations exist across populations. Notably, adiponectin levels, a key adipokine known for its insulin-sensitizing properties, are lower in Black/African populations compared to White/European populations [45, 46]. Furthermore, adiponectin has been independently linked to insulin clearance [46], providing insights into the biological mechanisms that may underlie population variations in IS. Second, we found that these proteins effectively tracked changes in IS in both BMI-adjusted and unadjusted models, with high concordance. This underscores their potential predictive values for monitoring changes in IS.

Our previous published work did not include instrumental variable analyses to infer causality of associations. With the expanded sample, we attempted to prioritize causal relationships among the 247 significant protein-IS associations observed in the meta-analysis using two-sample MR analysis. Eight candidate proteins showed evidence of causal effects on IS. Among these, Protein Tyrosine Phosphatase Receptor Type N2 (PTPRN2) is recognized as an autoantigen in insulin-dependent diabetes [47]. Additionally, increases in PTPRN2 protein levels have been shown to positively impact insulin secretion by increasing the number of dense core vesicles[48, 49]. While increased insulin secretion does not necessary equate to improved insulin sensitivity, our findings suggest that elevated PTPRN2 levels improve IS. This relationship warrants further investigation to clarify whether PTPRN2 influences IS directly or indirectly, possibly through mechanisms like insulin secretion regulation at different glucose levels.

Two candidate causal proteins are currently targets of drugs under investigation in registered clinical trials including Asialoglycoprotein Receptor 1 (ASGR1) and E-selectin (SELE). These drugs could ultimately demonstrate improvement in health outcomes through improved IS. For the former, our findings suggest lower levels of ASGR1 causally improved IS. This directionality is consistent with multiple lines of experimental and observation evidence demonstrating improved hepatic IS, reduced non-HDL cholesterol and triglycerides, and reduced risk of CVD in transformed human hepatocytes, mouse models, and population genetic studies of ASGR1 deficiency [50–54]. These insights also suggest that ASGR1 inhibitors such as Asiaglycorotein receptor 1 binding agent, initially explored for familial partial lipodystrophy treatment (ClinicalTrials.gov ID: NCT03514420), a condition often associated with IR due to abnormal fat distribution and impaired lipid as well as glucose metabolism [54] could potentially be repurposed for the treatment of IR. SELE is currently the target of several drugs under investigation in registered clinical trials. SELE is an indicator of endothelial dysfunction, a condition often present in metabolic diseases such as type 2 diabetes and cardiovascular diseases, both of which are characterized by IR [55]. Our finding suggests that increasing level of E-selectin reduces IS which aligns with multiple prior studies showing that serum SELE levels are inversely associated with IS [55–57] but evidence to support a causal association has generally been lacking. E-Selectin is a target for Selectin E antagonists, and several such antagonists such as Uproleselan, Rivipansel are in the third phase of clinical trials targeting the treatment of liver and kidney disease (ClinicalTrials.gov ID: NCT02871570, NCT02813798). Altogether, our findings suggest repurposing E-selectin antagonist to improve IS could improve cardiometabolic health.

In addition to PTPRN2 and SELE, we identified that Serine Hydroxymethyltransferase 1 (SHMT1), a key enzyme in serine and one-carbon metabolism, is linked to lower IS in our observational and MR analyses. Studies have identified SHMT1 as a novel biomarker for IR [58], and it has been found to be upregulated in diabetic mice[59]. Pharmacological inhibition of SHMT1/2 has been shown to increase circulating glycine levels[60], and dietary glycine supplementation has demonstrated benefits in improving insulin response, glucose tolerance and insulin sensitivity [61].

While PTPRN2, ASGR1, SELE and SHMT1 emerged as the most relevant proteins influencing IS, several other proteins such as MASP1, MVK, CES3, INHBC, and CDHR5 showed potential roles but had conflicting results between observational and MR analyses, warranting further investigation. Additionally, the tissue-specific roles of IL10RB in IR require additional investigation. More detailed discussion of these proteins and their potential effects on IS are included in the Supplementary Data.

A key strength of our study lies in its large-scale, multi-ethnic approach using gold standard measures of IS. The consistency between observational and interventional results, along with MR results, underscores the potential of these plasma proteins as therapeutic targets and biomarkers for IS. However, we acknowledge that circulating protein levels may not fully reflect the proteome in muscle cells or the pancreas, necessitating further research in these areas.

Additionally, we restricted profiling our intervention study participants to a small subset demonstrating a very substantial improvement in IS potentially limiting the generalizability of our findings. Future studies should examine a broader spectrum of changes in IS in both directions to provide a more comprehensive understanding of how plasma protein levels change with changes in IS. Further, a limitation of this study includes the composition of the sample population in the Stanford cohort, where 3.7% (38 out of 1,015) of the participants had diabetes. However, only 22 of 38 were undergoing treatment with antidiabetic medications and the medications used were not expected to affect the IS measurements, which may mitigate concerns regarding their potential confounding effects. While MR is a powerful tool for inferring causality, its effectiveness can be constrained by limited statistical power, particularly when the genetic instruments (cis-pQTLs) used explained only a small proportion of the variance in the plasma protein levels. Additionally, the robustness of the method may be compromised when the sample size for the gold standard measure of IS is low. Furthermore, our MR approach was performed on individuals of European descent only. Thus, the MR results may not be generalizable to other populations.

## Conclusion

Overall, our comprehensive integrative study of plasma proteins and genetic data offers insights into the molecular mechanisms of IS. Our study underscores the utility of these proteins as potential universal biomarkers for IS, serving not only as biomarkers but also as monitoring tools for changes in IS. Furthermore, our findings highlight the potential of these proteins as targets for drug development or repurposing. These insights contribute to the advancement of understanding and management of IR.

## Data Availability

All data produced in the present work are contained in the manuscript.

## Abbreviations

BMI: body mass index
CVD: cardiovascular disease
GWAS: genome-wide association studies
HEC: hyperinsulinemic-euglycemic clamp
IR: insulin resistance
IS: insulin sensitivity
IST: insulin suppression test
IVW: inverse variance weighted
MAF: minor allele frequency
MR: Mendelian Randomization
NPX: Normalized Protein eXpression
PEA: proximity extension assay
pQTL: protein quantitative trait loci
RCD: reduced-calorie diet
REML: Restricted Maximum Likelihood
RISC: the Relationship between Insulin Sensitivity and Cardiovascular Disease
SIRE: self-identified race/ethnicity
SSPG: steady-state plasma glucose
TZD: thiazolidinedione
ULSAM: Uppsala Longitudinal Study of Adult Men

## Declaration of competing interest

The authors declare no conflicts of interest.

CRediT authorship contribution statement:

PK, FA, and TLA designed the study. PK, NW, FA, TLA drafted and edited the manuscript, DZ, JWK, DJP, KTW, LS, LCL, SG, LL, and JRP reviewed and edited the manuscript. LL, JRP, and TLA contributed resources and supervised the project. TLA acquired fundings for the project. All authors approved the final version.

## References

[1] Reaven GM. Banting lecture 1988. Role of insulin resistance in human disease. Diabetes. 1988;37:1595–607.

[2] Reaven GM. Why Syndrome X? From Harold Himsworth to the insulin resistance syndrome. Cell Metab. 2005;1:9–14.

[3] Li M, Chi X, Wang Y, Setrerrahmane S, Xie W, Xu H. Trends in insulin resistance: insights into mechanisms and therapeutic strategy. Signal Transduct Target Ther. 2022;7:216.

[4] Park SY, Gautier JF, Chon S. Assessment of Insulin Secretion and Insulin Resistance in Human. Diabetes Metab J. 2021;45:641–54.

[5] Knowles JW, Assimes TL, Tsao PS, Natali A, Mari A, Quertermous T, et al. Measurement of insulin-mediated glucose uptake: direct comparison of the modified insulin suppression test and the euglycemic, hyperinsulinemic clamp. Metabolism: clinical and experimental. 2013;62:548–53.

[6] Katz A, Nambi SS, Mather K, Baron AD, Follmann DA, Sullivan G, et al. Quantitative insulin sensitivity check index: a simple, accurate method for assessing insulin sensitivity in humans. J Clin Endocrinol Metab. 2000;85:2402–10.

[7] Yeni-Komshian H, Carantoni M, Abbasi F, Reaven GM. Relationship between several surrogate estimates of insulin resistance and quantification of insulin-mediated glucose disposal in 490 healthy nondiabetic volunteers. Diabetes Care. 2000;23:171–5.

[8] Kim SH, Abbasi F, Reaven GM. Impact of degree of obesity on surrogate estimates of insulin resistance. Diabetes Care. 2004;27:1998–2002.

[9] Ingelsson E, Langenberg C, Hivert MF, Prokopenko I, Lyssenko V, Dupuis J, et al. Detailed physiologic characterization reveals diverse mechanisms for novel genetic Loci regulating glucose and insulin metabolism in humans. Diabetes. 2010;59:1266–75.

[10] De Souza AL, Batista GA, Alegre SM. Assessment of insulin sensitivity by the hyperinsulinemic euglycemic clamp: Comparison with the spectral analysis of photoplethysmography. J Diabetes Complications. 2017;31:128–33.

[11] Abbasi F, Silvers A, Viren J, Reaven GM. Relationship between several surrogate estimates of insulin resistance and a direct measure of insulin-mediated glucose disposal: Comparison of fasting versus post-glucose load measurements. Diabetes Res Clin Pract. 2018;136:108–15.

[12] Wik L, Nordberg N, Broberg J, Bjorkesten J, Assarsson E, Henriksson S, et al. Proximity Extension Assay in Combination with Next-Generation Sequencing for High-throughput Proteome-wide Analysis. Mol Cell Proteomics. 2021;20:100168.

[13] Zanetti D, Stell L, Gustafsson S, Abbasi F, Tsao PS, Knowles JW, et al. Plasma proteomic signatures of a direct measure of insulin sensitivity in two population cohorts. Diabetologia. 2023;66:1643–54.

[14] Hedstrand H. A study of middle-aged men with particular reference to risk factors for cardiovascular disease. Ups J Med Sci Suppl. 1975;19:1–61.

[15] Hills SA, Balkau B, Coppack SW, Dekker JM, Mari A, Natali A, et al. The EGIR-RISC STUDY (The European group for the study of insulin resistance: relationship between insulin sensitivity and cardiovascular disease risk): I. Methodology and objectives. Diabetologia. 2004;47:566–70.

[16] Ingelsson E, Sundstrom J, Arnlov J, Zethelius B, Lind L. Insulin resistance and risk of congestive heart failure. JAMA. 2005;294:334–41.

[17] Wiberg B, Sundstrom J, Zethelius B, Lind L. Insulin sensitivity measured by the euglycaemic insulin clamp and proinsulin levels as predictors of stroke in elderly men. Diabetologia. 2009;52:90–6.

[18] Ryan MC, Fenster Farin HM, Abbasi F, Reaven GM. Comparison of waist circumference versus body mass index in diagnosing metabolic syndrome and identifying apparently healthy subjects at increased risk of cardiovascular disease. Am J Cardiol. 2008;102:40–6.

[19] Raygor V, Abbasi F, Lazzeroni LC, Kim S, Ingelsson E, Reaven GM, et al. Impact of race/ethnicity on insulin resistance and hypertriglyceridaemia. Diab Vasc Dis Res. 2019;16:153–9.

[20] Abbasi F, Lamendola C, Harris CS, Harris V, Tsai MS, Tripathi P, et al. Statins Are Associated With Increased Insulin Resistance and Secretion. Arterioscler Thromb Vasc Biol. 2021;41:2786–97.

[21] Abbasi F, Robakis TK, Myoraku A, Watson KT, Wroolie T, Rasgon NL. Insulin resistance and accelerated cognitive aging. Psychoneuroendocrinology. 2023;147:105944.

[22] Abbasi F, Chen YD, Farin HM, Lamendola C, Reaven GM. Comparison of three treatment approaches to decreasing cardiovascular disease risk in nondiabetic insulin-resistant dyslipidemic subjects. Am J Cardiol. 2008;102:64–9.

[23] Abbasi F, Farin HM, Lamendola C, McGraw L, McLaughlin T, Reaven GM. Pioglitazone administration decreases cardiovascular disease risk factors in insulin-resistant smokers. Metabolism: clinical and experimental. 2008;57:1108–14.

[24] McLaughlin TM, Liu T, Yee G, Abbasi F, Lamendola C, Reaven GM, et al. Pioglitazone increases the proportion of small cells in human abdominal subcutaneous adipose tissue. Obesity (Silver Spring). 2010;18:926–31.

[25] McLaughlin T, Carter S, Lamendola C, Abbasi F, Yee G, Schaaf P, et al. Effects of moderate variations in macronutrient composition on weight loss and reduction in cardiovascular disease risk in obese, insulin-resistant adults. Am J Clin Nutr. 2006;84:813–21.

[26] Kim SH, Abbasi F, Lamendola C, Liu A, Ariel D, Schaaf P, et al. Benefits of liraglutide treatment in overweight and obese older individuals with prediabetes. Diabetes Care. 2013;36:3276–82.

[27] DeFronzo RA, Tobin JD, Andres R. Glucose clamp technique: a method for quantifying insulin secretion and resistance. Am J Physiol. 1979;237:E214–23.

[28] Pei D, Jones CN, Bhargava R, Chen YD, Reaven GM. Evaluation of octreotide to assess insulin-mediated glucose disposal by the insulin suppression test. Diabetologia. 1994;37:843–5.

[29] Shen SW, Reaven GM, Farquhar JW. Comparison of impedance to insulin-mediated glucose uptake in normal subjects and in subjects with latent diabetes. J Clin Invest. 1970;49:2151–60.

[30] Harano Y, Hidaka H, Takatsuki K, Ohgaku S, Haneda M, Motoi S, et al. Glucose, insulin, and somatostatin infusion for the determination of insulin sensitivity in vivo. Metabolism: clinical and experimental. 1978;27:1449–52.

[31] Otten J, Ahren B, Olsson T. Surrogate measures of insulin sensitivity vs the hyperinsulinaemic-euglycaemic clamp: a meta-analysis. Diabetologia. 2014;57:1781–8.

[32] Petrie JR. Evidence-based estimation of insulin resistance. Diabetologia. 2014;57:1743–5.

[33] Greenfield MS, Doberne L, Kraemer F, Tobey T, Reaven G. Assessment of insulin resistance with the insulin suppression test and the euglycemic clamp. Diabetes. 1981;30:387–92.

[34] Fuchsberger C, Abecasis GR, Hinds DA. minimac2: faster genotype imputation. Bioinformatics. 2015;31:782–4.

[35] Das S, Forer L, Schonherr S, Sidore C, Locke AE, Kwong A, et al. Next-generation genotype imputation service and methods. Nat Genet. 2016;48:1284–7.

[36] Taliun D, Harris DN, Kessler MD, Carlson J, Szpiech ZA, Torres R, et al. Sequencing of 53,831 diverse genomes from the NHLBI TOPMed Program. Nature. 2021;590:290–9.

[37] Knowles JW, Xie W, Zhang Z, Chennamsetty I, Assimes TL, Paananen J, et al. Identification and validation of N-acetyltransferase 2 as an insulin sensitivity gene. J Clin Invest. 2015;125:1739–51.

[38] Viechtbauer W. Conducting Meta-Analyses in R with the metafor Package. Journal of Statistical Software. 2010;36:1–48.

[39] Hemani G, Zheng J, Elsworth B, Wade KH, Haberland V, Baird D, et al. The MR-Base platform supports systematic causal inference across the human phenome. Elife. 2018;7.

[40] Mbatchou J, Barnard L, Backman J, Marcketta A, Kosmicki JA, Ziyatdinov A, et al. Computationally efficient whole-genome regression for quantitative and binary traits. Nat Genet. 2021;53:1097–103.

[41] Willer CJ, Li Y, Abecasis GR. METAL: fast and efficient meta-analysis of genomewide association scans. Bioinformatics. 2010;26:2190–1.

[42] Sun BB, Chiou J, Traylor M, Benner C, Hsu Y-H, Richardson TG, et al. Genetic regulation of the human plasma proteome in 54,306 UK Biobank participants. bioRxiv. 2022:2022.06.17.496443.

[43] Genomes Project C, Auton A, Brooks LD, Durbin RM, Garrison EP, Kang HM, et al. A global reference for human genetic variation. Nature. 2015;526:68–74.

[44] Yarmolinsky J, Bonilla C, Haycock PC, Langdon RJQ, Lotta LA, Langenberg C, et al. Circulating Selenium and Prostate Cancer Risk: A Mendelian Randomization Analysis. J Natl Cancer Inst. 2018;110:1035–8.

[45] Hakim O, Bello O, Ladwa M, Shojaee-Moradie F, Jackson N, Peacock JL, et al. Adiponectin is associated with insulin sensitivity in white European men but not black African men. Diabet Med. 2021;38:e14571.

[46] Ladwa M, Bello O, Hakim O, Boselli ML, Shojaee-Moradie F, Umpleby AM, et al. Exploring the determinants of ethnic differences in insulin clearance between men of Black African and White European ethnicity. Acta Diabetol. 2022;59:329–37.

[47] Kelleher KJ, Sheils TK, Mathias SL, Yang JJ, Metzger VT, Siramshetty VB, et al. Pharos 2023: an integrated resource for the understudied human proteome. Nucleic Acids Res. 2023;51:D1405–D16.

[48] Carmona GN, Nishimura T, Schindler CW, Panlilio LV, Notkins AL. The dense core vesicle protein IA-2, but not IA-2beta, is required for active avoidance learning. Neuroscience. 2014;269:35–42.

[49] Bian X, Wasserfall C, Wallstrom G, Wang J, Wang H, Barker K, et al. Tracking the Antibody Immunome in Type 1 Diabetes Using Protein Arrays. J Proteome Res. 2017;16:195–203.

[50] Nioi P, Sigurdsson A, Thorleifsson G, Helgason H, Agustsdottir AB, Norddahl GL, et al. Variant ASGR1 Associated with a Reduced Risk of Coronary Artery Disease. N Engl J Med. 2016;374:2131–41.

[51] Xie B, Shi X, Li Y, Xia B, Zhou J, Du M, et al. Deficiency of ASGR1 in pigs recapitulates reduced risk factor for cardiovascular disease in humans. PLoS Genet. 2021;17:e1009891.

[52] Wang JQ, Li LL, Hu A, Deng G, Wei J, Li YF, et al. Inhibition of ASGR1 decreases lipid levels by promoting cholesterol excretion. Nature. 2022;608:413–20.

[53] Svecla M, Da Dalt L, Moregola A, Nour J, Baragetti A, Uboldi P, et al. ASGR1 deficiency diverts lipids toward adipose tissue but results in liver damage during obesity. Cardiovasc Diabetol. 2024;23:42.

[54] Yu X, Tao J, Wu Y, Chen Y, Li P, Yang F, et al. Deficiency of ASGR1 Alleviates Diet-Induced Systemic Insulin Resistance via Improved Hepatic Insulin Sensitivity. Diabetes Metab J. 2024;48:802–15.

[55] Adamska A, Karczewska-Kupczewska M, Nikolajuk A, Otziomek E, Gorska M, Kowalska I, et al. Relationships of serum soluble E-selectin concentration with insulin sensitivity and metabolic flexibility in lean and obese women. Endocrine. 2014;45:422–9.

[56] Taniguchi A, Fukushima M, Nakai Y, Kuroe A, Yamano G, Yanagawa T, et al. Soluble E-selectin, leptin, triglycerides, and insulin resistance in nonobese Japanese type 2 diabetic patients. Metabolism: clinical and experimental. 2005;54:376–80.

[57] Delibasi T, Karbek B, Bozkurt NC, Cakir E, Gungunes A, Unsal OO, et al. Circulating E-selectin levels and insulin resistance are associated with early stages of atherosclerosis in nonfunctional adrenal incidentaloma. Arch Endocrinol Metab. 2015;59:310–7.

[58] Pujar MK, Vastrad B, Vastrad C. Integrative Analyses of Genes Associated with Subcutaneous Insulin Resistance. Biomolecules. 2019;9.

[59] Handzlik MK, Gengatharan JM, Frizzi KE, McGregor GH, Martino C, Rahman G, et al. Insulin-regulated serine and lipid metabolism drive peripheral neuropathy. Nature. 2023;614:118–24.

[60] McBride MJ, Hunter CJ, Zhang Z, TeSlaa T, Xu X, Ducker GS, et al. Glycine homeostasis requires reverse SHMT flux. Cell Metab. 2024;36:103–15 e4.

[61] Alves A, Bassot A, Bulteau AL, Pirola L, Morio B. Glycine Metabolism and Its Alterations in Obesity and Metabolic Diseases. Nutrients. 2019;11.

