## Supplementary data for "Cross-sectional, interventional, and causal investigation of insulin sensitivity using plasma proteomics in diverse populations"

Methods

*Proteomics profiling*

In total, 1,144 plasma samples were sent for proteomic profiling, including a subset with both pre- and post-interventions. One sample. was excluded due to a low count (<500) across all panel. As part of our own quality control (QC), we identified outliers within each protein panel by principal component analysis, and assessed median and inter-quartile range values for normalized NPX across protein samples. Data points were excluded if they exhibited a standardized PC1 or PC2 value more than five standard deviations from the mean (in standardized PCA, this mean is zero) or if they had a median NPX or IQR of NPX more than five standard deviations from their respective mean values. This analysis was performed using the “OlinkAnalyze” package in R, which is developed and maintained by Olink Proteomics Data Science Team. After removing outliers and excluding data points with QC or assay warnings, we retained 1,134 participants and 1,470 proteins for analysis.

*Genotyping*

DNA extraction services were provided by North American Genomics and genotyping was conducted at Thermo Fisher Scientific’s laboratory in Santa Clara, CA. After excluding three samples due to inadequate aliquot concentrations, 937 samples were retained for analysis.

For quality control, we removed samples with gender mismatches or those exceeding 6 standard deviations from the 1000 Genomes European mean. Prior to imputation, variants were filtered to exclude A/T or C/G alleles with minor allele frequency (MAF) exceeding 0.4, alleles mismatches with the reference panel, significant allele frequency discrepancies exceeding 0.2 with the reference panel, or if the SNPs are absent in the reference panel. Imputation was performed using the TOPMed reference panel [1-3] with allele frequencies showing a strong correlation with the reference panel (r^2^ = 0.98).

Genotyping and imputation for the RISC and ULSAM cohorts were described in detail elsewhere [4]. Briefly, RISC used the Affymetrix 6.0 microarray, and ULSAM used the Illumina Omni2.5M array, both were imputed to the 1000 Genome Project reference panel. We applied the same QC as that applied to the Stanford cohort.

Discussion

Our analysis also uncovered the role of MASP1 and MVK in insulin sensitivity. MBL Associated Serine Protease 1 (MASP1), known for its positive association with fasting glucose levels, 2-hour glucose levels, prediabetes, and diabetes [5-8]. This contradicts the positive causal relationship between MASP1 and insulin sensitivity suggested by our MR analysis. Similarly, Mevalonate Kinase (MVK) is primarily involved in cholesterol biosynthesis [9] but it has not explicitly linked to insulin sensitivity. Its role in metabolic pathways suggests possible indirect effects on insulin sensitivity. However, the conflicting findings from observational and MR analysis for both MASP1 and MVK underscore the necessity of further research to elucidate the role of these proteins in insulin sensitivity.

In addition to these findings, we identified Cadherin-Related Family Member 5 (CDHR5), which has limited evidence connecting it to insulin sensitivity. CDHR5 plays a critical role in cell adhesion and the branching of organs under normal physiological conditions[10]. In our study, there is conflicting evidence between the observational and MR analyses, despite the cis-pQTLs used as instrumental variables in the MR analysis explained a substantial amount of variance (11.34%). Further studies are required to resolve these inconsistencies.

Similarly, conflicting evidence between observational and MR analyses was also observed for Carboxylesterase 3 (CES). CES3 is involved in lipid metabolism, primarily through the hydrolysis of triglycerides in adipose tissue and the liver. Ablation of CES3 expression has been shown to lower blood lipid levels and improve glucose tolerance in mice, suggesting a protective role against insulin resistance [11-13]. However, our study presents contrasting evidence with observational analysis showing inverse relation and MR analysis showing positive relation. Additionally, the instrumental variables used in MR analysis explaining only 2.36% of the variance in plasma CES3 levels. This limited evidence of causal inference, combined with CES3 being measured only in the Olink Explore panel used in Stanford studies, underscores the need for further studies that measure CES3 levels and gold standard measures of insulin sensitivity to clarify the association.

Additionally, another candidate protein influencing insulin sensitivity is Interleukin 10 Receptor Subunit Beta (IL10RB) is part of the IL10 receptor complex and plays a crucial role in anti-inflammatory signaling. However, the role of IL10 in insulin resistance is complex and context-dependent. In human white adipose tissue, IL10 is upregulated in pro-inflammatory macrophages of obese and insulin resistant individuals, even though it does not directly affect adipocytes [14]. In contrast, in the liver, IL10 has a protective role in diet-induced insulin resistance by reducing inflammation and inhibiting gluconeogenesis [15]. Interestingly, IL10 deficiency in mice increased energy expenditure and protected against diet-induced obesity [16]. These contrasting roles of IL10 in different tissues suggest the need for further investigation into the tissue-specific role of IL10RB in insulin sensitivity.

Our analysis also highlighted Inhibin Subunit Beta C (INHBC), part of the TGF-beta family, regulates follicle-stimulating hormone and insulin secretion [17], and is highly expressed in the liver [18]. While our observational analysis found an inverse relationship between INHBC and insulin sensitivity, our MR analysis showed a positive association, with 5.9% of the variance explained by instrumental variables. Previous MR study linked INHBC to dyslipidemia, coronary artery disease, and non-alcoholic fatty liver disease [19]. Additionally, INHBC promotes pro-inflammatory cytokines such as IL6 and TNF-alpha in diabetic nephropathy [20]. A high-throughput proteomic study also associated elevated INHBC with increased risk of incident type 2 diabetes and lower insulin sensitivity [21], supporting our observational findings. This reinforces the need for further research and additional instrumental variables to strengthen the causal inference of INHBC’s role in insulin sensitivity.

References for Supplementary File

[1] Fuchsberger C, Abecasis GR, Hinds DA. minimac2: faster genotype imputation. Bioinformatics. 2015;31:782-4.

[2] Das S, Forer L, Schonherr S, Sidore C, Locke AE, Kwong A, et al. Next-generation genotype imputation service and methods. Nat Genet. 2016;48:1284-7.

[3] Taliun D, Harris DN, Kessler MD, Carlson J, Szpiech ZA, Torres R, et al. Sequencing of 53,831 diverse genomes from the NHLBI TOPMed Program. Nature. 2021;590:290-9.

[4] Knowles JW, Xie W, Zhang Z, Chennamsetty I, Assimes TL, Paananen J, et al. Identification and validation of N-acetyltransferase 2 as an insulin sensitivity gene. J Clin Invest. 2016;126:403.

[5] Jenny L, Ajjan R, King R, Thiel S, Schroeder V. Plasma levels of mannan-binding lectin-associated serine proteases MASP-1 and MASP-2 are elevated in type 1 diabetes and correlate with glycaemic control. Clin Exp Immunol. 2015;180:227-32.

[6] von Toerne C, Huth C, de Las Heras Gala T, Kronenberg F, Herder C, Koenig W, et al. MASP1, THBS1, GPLD1 and ApoA-IV are novel biomarkers associated with prediabetes: the KORA F4 study. Diabetologia. 2016;59:1882-92.

[7] Krogh SS, Holt CB, Steffensen R, Funck KL, Hoyem P, Laugesen E, et al. Plasma levels of MASP-1, MASP-3 and MAp44 in patients with type 2 diabetes: influence of glycaemic control, body composition and polymorphisms in the MASP1 gene. Clin Exp Immunol. 2017;189:103-12.

[8] Kietsiriroje N, Scott GE, Ajjan RA, Broz J, Schroeder V, Campbell MD. Plasma levels of mannan-binding lectin-associated serine proteases are increased in type 1 diabetes patients with insulin resistance. Clin Exp Immunol. 2024;215:58-64.

[9] Junyent M, Parnell LD, Lai CQ, Lee YC, Smith CE, Arnett DK, et al. Novel variants at KCTD10, MVK, and MMAB genes interact with dietary carbohydrates to modulate HDL-cholesterol concentrations in the Genetics of Lipid Lowering Drugs and Diet Network Study. Am J Clin Nutr. 2009;90:686-94.

[10] Gao J, Wang M, Li T, Liu Q, You L, Liao Q. Up-regulation of CDHR5 expression promotes malignant phenotype of pancreatic ductal adenocarcinoma. J Cell Mol Med. 2020;24:12726-35.

[11] Wei E, Ben Ali Y, Lyon J, Wang H, Nelson R, Dolinsky VW, et al. Loss of TGH/Ces3 in mice decreases blood lipids, improves glucose tolerance, and increases energy expenditure. Cell Metab. 2010;11:183-93.

[12] Lian J, Quiroga AD, Li L, Lehner R. Ces3/TGH deficiency improves dyslipidemia and reduces atherosclerosis in Ldlr(-/-) mice. Circ Res. 2012;111:982-90.

[13] Lian J, Wei E, Wang SP, Quiroga AD, Li L, Di Pardo A, et al. Liver specific inactivation of carboxylesterase 3/triacylglycerol hydrolase decreases blood lipids without causing severe steatosis in mice. Hepatology. 2012;56:2154-62.

[14] Acosta JR, Tavira B, Douagi I, Kulyte A, Arner P, Ryden M, et al. Human-Specific Function of IL-10 in Adipose Tissue Linked to Insulin Resistance. J Clin Endocrinol Metab. 2019;104:4552-62.

[15] Cintra DE, Pauli JR, Araujo EP, Moraes JC, de Souza CT, Milanski M, et al. Interleukin-10 is a protective factor against diet-induced insulin resistance in liver. J Hepatol. 2008;48:628-37.

[16] Rajbhandari P, Thomas BJ, Feng AC, Hong C, Wang J, Vergnes L, et al. IL-10 Signaling Remodels Adipose Chromatin Architecture to Limit Thermogenesis and Energy Expenditure. Cell. 2018;172:218-33 e17.

[17] Namwanje M, Brown CW. Activins and Inhibins: Roles in Development, Physiology, and Disease. Cold Spring Harb Perspect Biol. 2016;8.

[18] Fagerberg L, Hallstrom BM, Oksvold P, Kampf C, Djureinovic D, Odeberg J, et al. Analysis of the human tissue-specific expression by genome-wide integration of transcriptomics and antibody-based proteomics. Mol Cell Proteomics. 2014;13:397-406.

[19] Loh NY, Rosoff DB, Richmond R, Noordam R, Smith GD, Ray D, et al. Bidirectional Mendelian randomization highlights causal relationships between circulating INHBC and multiple cardiometabolic diseases and traits. Diabetes. 2024.

[20] Du XY, Zheng BT, Pang Y, Zhang W, Liu M, Xu XL, et al. The potential mechanism of INHBC and CSF1R in diabetic nephropathy. Eur Rev Med Pharmacol Sci. 2020;24:1970-8.

[21] Cronje HT, Mi MY, Austin TR, Biggs ML, Siscovick DS, Lemaitre RN, et al. Plasma Proteomic Risk Markers of Incident Type 2 Diabetes Reflect Physiologically Distinct Components of Glucose-Insulin Homeostasis. Diabetes. 2023;72:666-73.
